# Knowledge and awareness of Pre-exposure Prophylaxis among men in sub-Saharan Africa: A scoping review protocol

**DOI:** 10.1101/2023.03.09.23287056

**Authors:** Oluwaseun Abdulganiyu Badru, Mbuzeleni Hlongwa, Oluwafemi Atanda Adeagbo

## Abstract

**Introduction:** About 38.4 million people were living with HIV as of 2021, and HIV is more prevalent in sub-Saharan Africa. Pre-exposure prophylaxis (PrEP) is highly effective in HIV prevention. Despite the efficacy of PrEP, many persons, including men, do not have adequate knowledge and awareness of PrEP, and reviews on knowledge and awareness among men are scarce. This review aims to assess and synthesize the knowledge and awareness of PrEP among persons assigned as males at birth in sub-Saharan Africa.

**Method and analysis:** The proposed scoping review will be conducted in accordance with the PRISMA Extension for Scoping Reviews (PRISMA-ScR): Checklist and Explanation. The following information sources will be searched to retrieve relevant studies for this review: CINAHL, MEDLINE (Ovid), PubMed, SCOPUS, and Web of Science. Google Scholar, The Union Catalogue of Theses and Dissertations (UCTD) and SA ePublications via SABINET Online, WorldCat Dissertations and Theses via OCLC, ResearchGate, and American Doctoral Dissertations via EBSCOhost. All study designs, except existing reviews, will be included. All screenings (abstract screening and full-text screening) and data extraction will be conducted independently by two reviewers. Quantitative findings will be presented with frequency and percentages, while qualitative thematic analysis will be used to analyze qualitative findings.

**Ethics and dissemination:** Due to the use of primary published data, ethical approval is not required for this review. The results of this review will be presented at a conference and published in a high-impact peer-review journal.

## INTRODUCTION

About 38.4 million people were living with HIV as of 2021, which is more prevalent in sub-Saharan Africa [1]. Abstinence, appropriate condom use, and biomedical prevention such as post-exposure prophylaxis (PEP) and pre-exposure prophylaxis (PrEP) are strategies to prevent HIV [2]. In 2012, the World Health Organization (WHO) recommended PrEP to priority groups, such as transgender people, sex workers (SWs), and men who have sex with other men (MSM) [3]. Later, in 2015, the WHO recommended PrEP to persons with a substantial risk of HIV [4].

PrEP, a biomedical prevention intervention, has provided the opportunity to limit HIV incidence among priority groups and the general population [5]. PrEP is highly effective in HIV prevention [6–8]. Daily use of PrEP reduces HIV risk by 75% to 99% [9,10]. However, the low perceived risk of HIV and poor awareness and knowledge of PrEP limits the use of PrEP, especially in HIV hyperendemic settings [10,11].

Despite the efficacy of PrEP, many persons, including men (defined as those assigned male at birth: cisgender men, MSM and transgender women), at risk of HIV do not have adequate knowledge or are not aware of PrEP. For example, only 45% of transgender women surveyed in South Africa were aware of PrEP [12]. Similarly, only 45.8% of MSM and transgender women were aware of PrEP in Zimbabwe [9], and more than half (53.6%) of MSM in Nigeria were aware of PrEP in Nigeria [13].

Several individual and interpersonal factors are associated with awareness and knowledge of PrEP among men; individual factors such as self-efficacy, having health insurance and residing in an urban area determine awareness and knowledge of PrEP [14,15]. Interpersonal factors such as having social support and being a member of a lesbian, gay, bisexual and transgender organization, and having a partner who is living with HIV are associated with PrEP awareness [13–15].

Generally, men are less likely to know their HIV status and are more likely to be unengaged in HIV care than women [16]. For instance, 90% of women in South Africa are aware of their HIV status compared to 82% of men, and more women are on antiretroviral therapy than men (65% versus 54%) as of the end of 2016 [16,17]. This suggests that men are behind in seeking care HIV care and prevention, due to multiple interpersonal and structural factors, including long waiting hours at the clinics, stigma, financial constraints due to travelling costs, discrimination, privacy concerns, poor knowledge of PrEP and where to assess it [18,19].

HIV treatment as prevention is effective in the prevention of HIV transmission from a seropositive person to an uninfected person if used consistently and as prescribed [20]. However, prevention is often said to be better than treatment, and the use of PrEP may be considered ‘HIV prevention as treatment’, especially for MSM and transgender women, as they have a higher risk of contracting HIV [1].

Earlier reviews on PrEP have focused on willingness, uptake, adherence, barriers, and facilitators to PrEP among MSM [21–25], while one earlier review investigated PEP awareness among MSM [26]. The reason for the focus on MSM is understandable, as MSM are disproportionately more likely at risk of HIV, and the initial focus of PrEP was on priority groups, including transgender persons and MSM [3]. Kamitani *et al*. [22], in their scoping review, found that most studies on PrEP focused on MSM; the recommendation of PrEP for MSM by the WHO may have contributed to the focus on MSM [27]. However, not many primary studies have investigated the knowledge and awareness of PrEP among the general population or heterosexual men only [27]. By extension, reviews on knowledge and awareness of PrEP are scarce. To bridge this gap, this review aims to assess and synthesize the knowledge and awareness of PrEP among persons assigned as males at birth in sub-Saharan Africa.

### Objectives

- To assess the knowledge and awareness of PrEP among men in sub-Saharan Africa.
- To identify factors associated with knowledge and awareness of PrEP among men in sub-Saharan Africa.

## METHODS AND ANALYSIS

The proposed scoping review will be conducted following the PRISMA Extension for Scoping Reviews (PRISMA-ScR): Checklist and Explanation [28].

1. Identifying the research questions,
2. Identifying relevant studies,
3. Study selection,
4. Charting the data,
5. Collating, summarizing, and reporting results.

### Research questions

- Are men in sub-Saharan Africa knowledgeable and aware of PrEP?
- What are the associated factors with knowledge and awareness of PrEP among men in sub-Saharan Africa?

### Identifying relevant studies

The search strategy aims to find both published and unpublished studies. We will conduct a preliminary search on PubMed, after which we will analyze text words and index terms used to describe the articles. This will assist in the creation of a search strategy that is specific to each information source. The following information sources will be searched to retrieve relevant studies for this review: CINAHL, MEDLINE (Ovid), PubMed, SCOPUS, and Web of Science. Google Scholar, The Union Catalogue of Theses and Dissertations (UCTD) and SA ePublications via SABINET Online, WorldCat Dissertations and Theses via OCLC, ResearchGate, and American Doctoral Dissertations via EBSCOhost will all be searched for unpublished academic research (theses and/or dissertations). All studies eligible for inclusion will have their reference lists checked for potential additional articles. Our search technique will include boolean terms (AND, OR) and Medical Subject Headings (MeSH) phrases. The following key search words will be used: knowledge, awareness, pre-exposure prophylaxis, men, and sub-Saharan Africa. Some relevant MeSH terms include “Health Knowledge, Attitudes, Practice”[Mesh], “Awareness”[Mesh], “Pre-Exposure Prophylaxis”[Mesh], “Men”[Mesh], and “Africa South of the Sahara”[Mesh]. Each keyword will be combined with relevant MeSH and synonymous terms with the “OR” Boolean operator to form a block, while each block will be combined with the “AND” Boolean operator (see S1 Table). We will also employ African country names and abbreviated terms like “east Africa/ west Africa” to ensure that articles indexed using African nation-specific names or regional terms are retrieved.

### Eligibility criteria Types of participants

Only studies focused on adults (18 years and above) assigned male at birth will be included.

### Type of outcome

Studies that assessed knowledge or awareness of PrEP among men in sub-Saharan Africa will be considered.

### Type of Studies

All study designs will be included in this scoping review, except for existing reviews (rapid review, scoping review, a systematic review with or without meta-analysis, among others). However, only the baseline findings of cohort studies and trials (randomized or non-randomized trials) will be reported. Furthermore, conference proceedings and relevant dissertations will be considered in this review.

### Other criteria

Studies published earlier than 2012 will be excluded since the first PrEP drug was approved in the year 2012 [29]. There will be no restriction on language.

### Study procedure and selection of the studies

All citations identified by the search will be downloaded and imported into Rayyan systematic review manager for deduplication and screening of articles [30]. Two authors (OAB and MH) will screen the title and abstract of all articles independently to determine their eligibility. Articles that do not meet the study’s eligibility criteria will be excluded. The full text of articles that appear to have met the eligibility criteria will be obtained and screened independently by two authors (OAA and OAB). Discussion or collaboration with a third reviewer will be used to settle any disputes between the two independent reviewers, if any.

### Charting the data

The Joanna Briggs Institute’s template for data extraction will be adapted for data extraction. The following data will be extracted from each study that meets the eligibility criteria: last name of the first author, year of publication, study objective(s), study setting, participant’s characteristics, sample size, the country where the study was conducted, sampling method, data collection method, data analysis, study limitations and implications, knowledge and/or awareness of PrEP (in the form of prevalence or responses to individual questions), factors associated with knowledge and awareness of PrEP (bivariate and multivariate analysis), and conclusions from authors. One author (OAB) will extract relevant data from all included studies, while two other authors (MH and OAA) will verify the extraction.

### Collating, summarizing, and reporting the results

Quantitative findings will be synthesized and summarized with a narrative approach. The prevalence of knowledge and awareness will be descriptively summarized. Furthermore, descriptive statistics will be used to represent the region based on the World Health Organization regional classification, sampling method (probability or non-probability), characteristics of the participants, and factors associated with knowledge and awareness of PrEP. Qualitative thematic analysis will be used to analyze qualitative studies. Depending on the findings, the quantitative and qualitative findings will be triangulated.

### Patient and public involvement

Patients or the public will not be directly involved in the design or dissemination of this review’s findings.

### Ethics and dissemination

Ethical clearance is not warranted as already published studies will be reviewed and synthesized. The findings of this review will be published in an open-access journal for adequate dissemination and publicity. The findings will be shared with relevant researchers, policymakers and non-governmental organizations interested in HIV prevention.

## DISCUSSION

This scoping review will assess the knowledge and awareness of PrEP and associated factors among men in sub-Saharan Africa. This review will be the first to chart the knowledge of men in sub-Saharan Africa. This review will help understand the level of PrEP knowledge among men in sub-Saharan Africa and the factors that predict PrEP knowledge. The identified predictors or determinants can serve as targets for future interventions to improve knowledge and awareness of PrEP among men. Furthermore, the findings of this review may lead to policy formation tailored towards men’s knowledge in several sub-Saharan African countries. The findings of this review will be disseminated in the form of conference presentations and peer-review publication.

## Data Availability

All relevant data are within the manuscript and its Supporting Information files.

